# Impact of COVID-19 pandemic on rare diseases- A case study on thalassaemia patients in Bangladesh

**DOI:** 10.1101/2021.03.29.21254589

**Authors:** Mohammad Sorowar Hossain, Farhana Runa, Abdullah Al Mosabbir

## Abstract

**Objectives:** Thalassaemia is a life-threatening rare disease, which requires regular blood transfusion and medical care. The information on how thalassaemia patients are affected during the unprecedented COVID-19 crisis is scarce. This study aimed to assess the impact of the COVID-19 pandemic on the blood transfusion and healthcare access of thalassaemia patients at the community level in Bangladesh.

**Methods:** A cross-sectional study was conducted among thalassaemia patients registered in a community-based ‘ thalassaemia registry’ in Jamapur, Bangladesh.

**Results:** As compared to pre-COVID-19 time, the number of blood transfusions among patients under the thalassaemia registry was significantly reduced during COVID-19 pandemic (190 units versus 81 units). In addition, the median number of red cell transfusions per patient was dropped significantly from 4 units to one unit. Over 80% of patient had no access to healthcare services at all during the early phase of the pandemic.

**Conclusions:** Emergency response with appropriate mitigative measures must be a priority for addressing an acute shortage of blood supply in situations like COVID-19 pandemic.

## Introduction

The unprecedented Coronavirus Disease 19 (COVID-19) pandemic ever faced by mankind has impacted every aspect of our life. Because of the chronic nature of the illness, individuals with rare diseases have to confront inadequacies of healthcare access and support even in regular times. Conceivably, the COVID-19 pandemic has made their lives more vulnerable and, perhaps, unbearable. The scenario is expected to be dire in the perspective of resource-poor countries. Rare diseases are often highly progressive, and life-threatening, arising largely from genetic causes. Globally, an estimated 300 million people suffer from 7000 rare diseases and 75% of them are children^1^.

Beta-thalassaemia, an inherited genetic disorder, is a well-known rare disease. This disease results from an absence and /or ineffective synthesis of beta-globin chains. Nearly 50,000 babies are born worldwide every year with a severe form of thalassaemia requiring regular blood transfusion. Once thought of as a localized disease in certain parts of the world, thalassaemias are becoming a global public health concern because of increasing international migration. In the Indian subcontinent, more than 200,000 thalassaemia patients (45–70 million carriers) need regular blood transfusion with chelation therapy ^2^. We present the case of thalassaemia to assess the impact of the COVID-19 pandemic on rare diseases in the South Asian perspective, particularly Bangladesh.

Bangladesh is a lower-middle-income country with a population of over 160 million. Despite a higher prevalence of thalassaemia (estimated 10-19 million carriers and 60,000–70,000 patients), knowledge about this disease is poor among general people. Besides, like other developing countries, access to healthcare is limited even for highly prioritized communicable diseases; therefore, necessary care for thalassaemia patients is not pursued with careful consideration.

The first official case of COVID-19 was declared on 8 March; the nationwide lockdown and related restrictions commenced on 24 March 2020. The pandemic took a toll on the healthcare system. Like other developing countries ^3^, Bangladesh faced a critical drop of blood supply especially during the early phase of COVID-19. How the nationwide lockdown and movement restrictions during the early phase of COVID-19 have affected the already miserable life of thalassaemia patients in a resource-poor country like Bangladesh needs thorough investigation for future policymaking. Therefore, we aimed to assess the impact of the COVID-19 pandemic on the management of blood transfusion and healthcare access of thalassaemia patients at the community level in Bangladesh.

## Methods

### Study setting and participants

This was a cross-sectional study conducted between 24 March 2020 and 30 July 2020 in Jamalpur district (one of the 64 administrative districts in Bangladesh) with a population of 2.3 million, situated 140□km northwest of the capital city, Dhaka). This study setting is representative of the majority part of Bangladesh in respective to demographic and socio-economic factors ^4^. After several years of research and community sensitization, Biomedical Research Foundation (BRF) took an initiative to develop a community-based ‘ thalassaemia registry’ in November 2019 in order to support thalassaemia patients and make awareness about this life-threatening yet preventable disease. As of date, 50 thalassemic patients have been registered. BRF has arranged financial support for 18 poor registered families to bear the cost of blood transfusion. It has also initiated a “blood for thalassaemia” drive to support these patients.

### Data collection and analysis

A short questionnaire was developed to assess the impact of the first four months (24 March-30 July 2020) of the COVID-19 pandemic. The survey questions aimed to assess the change in blood transfusion practice, impact on the overall health status of thalassaemic patients and access to healthcare service during the pandemic restrictions. Patients with thalassaemia or the parents of thalassaemic children (if age less than 18 years) were surveyed over the telephone. Demographic data were extracted from the ‘ thalassaemia registry’. To compare, transfusion data 16 weeks (December 2019-March 2020) before the commencement of COVID-19 were taken from the registry. Health status was grouped into 3 categories-mild weakness (weak but no problem doing the daily activity), moderate weakness (could do the daily activity but with difficulty), and severe weakness (bedridden). The study protocol was ethically approved by the Institutional Review Committee of the Biomedical Research Foundation (Memo: Ref. no: BRF/ERB/2020/003). Simple descriptive statistical analyses were used. SPSS Statistics software 22.0 (Armonk, NY: IBM Corp) was used to analyze data.

## Result

Out of 50 patients with thalassaemia registered in ‘ thalassaemia registry’, 4 patients died before the COVID-19 pandemic, 2 patients were transfusion-independent and 2 patients could not be reached over the phone. Finally, a total of 42 patients with transfusion-dependent thalassaemia (TDT) participated in the current study. The median age of the participants was 10.5 years (IQR 6.0-16.25) with slight male predominance (Table 1). More than 60% of the participants had HbE/beta thalassaemia and the rest had beta-thalassaemia major. Before COVID-19 pandemic, a total number of red cell transfusions was 190 units, whereas transfusion reduced more than half to 81 units during early phase. In addition, the median number of red cell transfusions per patient came down significantly to one unit during COVID-19 compared to four units before COVID-19 pandemic (p <0.001). The impact of this under transfusion was reflected on the health status of the patients. While the most of the patients (73.8%) reported mild weakness before COVID-19, majority complained moderate to severely weakness (38.1% and 28.6% respectively) during early pandemic making them unable to do daily activity or bedridden In addition, only 14.3% of the patients could consult a certified physician or hematologist. Among them one had to go to the capital city, Dhaka to avail the service. None of the patients and their family members reported to have COVID-19 infection during study period.

**Table 1.** Demographic characteristics, transfusion practice and health status of the patients

| Demographic characteristics (N= 42) |  |  |  |
| --- | --- | --- | --- |
| Age years |  |  |  |
| Median (IQR) | 10.5 (6.0-16.25) |  |  |
| Sex |  |  |  |
| Male | 22 (52.4%) |  |  |
| Female | 20 (47.6%) |  |  |
| Blood group |  |  |  |
| A | 13 (31.0%) |  |  |
| B | 13 (31.0%) |  |  |
| AB | 05 (11.9%) |  |  |
| O | 11 (26.2%) |  |  |
| Rh- | 01 (2.4%) |  |  |
| Diagnosis |  |  |  |
| HbE/beta thalassaemia | 26 (61.9%) |  |  |
| Beta Thalassaemia major | 16 (38.1%) |  |  |
| Transfusion practice |  |  |  |
|  | Before COVID-19 | During early COVID-19 | p |
| Total number of Red Cell Units transfused, n | 198 | 81 |  |
| Median number of Red Cell Units transfused/ person (IQR) | 4.0 (3.0-6.0) | 1.0 (0.0-4.0) | 0.000* |
| Health status |  |  |  |
| Mild weakness | 31 (73.8%) | 14 (33.3%) |  |
| Moderate weakness | 11 (26.2%) | 16 (38.1%) |  |
| Severe Weakness | 0 | 12 (28.6%) |  |
| Access to certified physician/Hematologist |  |  |  |
| Yes | 36 (85.7%) |  |  |
| No | 6 (14.3%) |  |  |

AbbreviationFs: IQR: interquartile range \*Wilcoxon rank-sum test was used; Two-tailed p value <0.05 was set as statistically significant.

## Discussion

In our study, we found most thalassaemic children were deprived of regular blood transfusion and medical follow-up at the district level in the time of COVID-19 restrictions. Over 80% thalassemic children had no access to healthcare services at all. A survey conducted by EURODIS (a big network of rare diseases) in 36 developed countries also found 84% of people living with a rare disease experienced disruption of care due to the COVID-19 pandemic ^5^. In the early days of the pandemic, mass panic, stigmatization, fear of uncertainty and infection caused shutdown of most public and private hospitals/clinics, except few specialized COVID-19 hospitals in Bangladesh. In some incidences, medical doctors also stayed away from hospitals due to limited supply of personal protective equipment (PPE) and fear of contracting an infection ^6–8^. From July 2020, the situation started to improve due to relaxation of lockdown and reduction of mass panic.

For survival, thalassaemia patients require regular blood transfusion, chelation therapy and monitoring of health status under specialized medical expertise, especially by hematologists. Notably, most specialized thalassaemia screening and treatment centers are located in large cities, mainly in Dhaka, the capital city of Bangladesh. Therefore, most patients residing in rural community are marginalized and deprived of proper care due to a severe lack of specialized doctors and unavailability of chelating drugs. So, patients, by no choice, have to rely on general practitioners in some cases to pediatricians where available. Moreover, like other resource-poor countries^9^, thalassaemia patients are generally under-transfused in Bangladesh. A study on Southeast Asian countries including Bangladesh reported a 38% shortfall of blood supplies^3^. In Bangladesh, voluntary non-remunerated donations only contribute 31% of the need while 60% donations come from friends and relatives ^10^. Consequently, the pandemic caused a near shutdown in blood supply due to nationwide lockdown, panic, travel restriction, cessation of blood donation camps. The life of patients with TDT, who rely on regular blood transfusion for survival, got severely impacted. As a result, about 30% of patients who had either mild or moderate weakness before the pandemic became severely weak and bed bound.

The major limitation of this study is the small sample size. However, this study has been conducted in a rural area using a community-based thalassaemia registry. Therefore, this study might represent the challenges faced by most, if not all, patients with thalassaemia in Bangladesh as more than 70% of people live in rural areas.

Even before the COVID-19 pandemic, the lives of thalassemic children were precarious, particularly in developing countries including Bangladesh with a fragile healthcare system. The COVID-19 pandemic has exacerbated this situation further due to an acute shortage of blood supply and inadequate medical care. It is, therefore, an important call to attention to the policymakers as well as international health care agencies like WHO regarding the needs of these patients and families with rare diseases in times of pandemics. Emergency response with appropriate mitigative measures must be a priority for ensuring an acute shortage of blood supply in situations like COVID-19 pandemic.

## Data Availability

The manuscript contains all available data

## Declaration of competing interest

Authors declare that there is no conflict of interest.

## Notes

### Competing Interest Statement

The authors have declared no competing interest.

### Funding Statement

Not applicable

### Author Declarations

The study protocol was ethically approved by the Institutional Review Committee of the Biomedical Research Foundation (Memo: Ref. no: BRF/ERB/2020/003).

